# GABAergic modulation of beta power enhances motor adaptation in frontotemporal lobar degeneration

**DOI:** 10.1101/2024.06.28.24309636

**Authors:** Laura E. Hughes, Natalie E. Adams, Matthew A. Rouse, Michelle Naessens, Alexander Shaw, Alexander G. Murley, Thomas E. Cope, Negin Holland, David Nesbitt, Duncan Street, David J. Whiteside, James B. Rowe

## Abstract

The impairment of behavioural control is a characteristic feature of disorders associated with frontotemporal lobar degeneration (FTLD). Behavioural disinhibition and impulsivity in these disorders are linked to abnormal neurophysiology of the frontal lobe, such as the loss beta-band power and changes in prefrontal GABAergic neurotransmission. Here we test the hypothesis that a pharmacological increase of GABA would concurrently improve cortical beta-band power and adaptive behavioural control in people with behavioural-variant frontotemporal dementia (bvFTD), and progressive supranuclear palsy (PSP, Richardson’s syndrome). We recorded magnetoencephalography during a visuomotor task that measures participants’ ability to adapt motor responses to visual feedback. Tiagabine, a GABA re-uptake inhibitor, was used as a pharmacological probe in a double-blind placebo controlled crossover design. The study included 11 people with bvFTD, 11 people with PSP and 20 healthy age-matched controls. Behavioural performance and beta power were examined with linear mixed models examined changes in, to estimate motor learning over time and the response to tiagabine. Significant beta power differences were source-localised using linear-constraint minimum variance beamformer. As predicted, participants with bvFTD and PSP were impaired behaviourally, and the beta power associated with movement, learning and accuracy, was diminished compared to controls. Tiagabine facilitated partial recovery of the impairments in behaviour and beta power over trials, moderated by executive function, such that the greatest improvements were seen in those with higher cognitive scores. The beamformer localised the physiological effects of disease and tiagabine treatment to frontal cortices, and confirmed the right prefrontal cortex as a key site of drug by group interaction. We interpret the differential response to tiagabine between bvFTD and PSP as a function of baseline differences in atrophy and physiology. In summary, behavioural and neurophysiological deficits can be mitigated by enhancement of GABAergic neurotransmission. Clinical trials are warranted to test for enduring clinical benefits from this restorative-psychopharmacology strategy.

## Introduction

The behavioural features of disorders associated with frontotemporal lobar degeneration (FTLD), include impulsivity and disinhibition with cognitive and motor inflexibility. They have a severe consequence with earlier loss of independence, higher carer burden, and reduced survival, highlighting the urgent need for symptomatic treatments.^1, 2^ For the development of potential therapeutic strategies, restoration of pharmacological and neurophysiological functions is more tractable then reversal of atrophy. The case for symptomatic treatment would be supported where neurochemical modulation leads to measurable changes in neurophysiology and behaviour.

While there are currently no disease modifying treatments for FTLD-associated disorders, small pharmacological trials have addressed the loss of critical neurotransmitters,^3, 4^ with variable results^5^. For example, pharmacological trials modifying serotonergic transmission have shown promising effects on behaviour and may be of clinical benefit.^6–8^ In some people with progressive supranuclear palsy (PSP) motor and behavioural impairments respond transiently to GABAergic modulation.^9, 10^ Trials of memantine, a non-competitive antagonist at NMDA and other receptors had no generalised efficacy,^11, 12^ but clinical responses may also depend on the severity of baseline deficits.^12^

We propose a strategic approach that links pharmacological modulation to behavioural effects via their underlying neurophysiological mechanisms, including physiological responses in prefrontal cortical circuits.^13^ The link between behaviour, neurophysiology and pharmacology may be observed in disease relevant changes in frequency-specific neuronal signals and oscillations. In FTLD-associated disorders, there is concordance across study designs in the loss of beta band connectivity in the resting-state, ^14, 15^ and loss of frontal beta power during task performance. ^16–19^

Neuronal signals in the beta frequency range (14-30Hz) are fundamental for the control of movement: a decrease in synchronous beta power occurs with movement planning and initiation (an event related desynchronization, ERD) and an increase above baseline is observed with movement cessation, (event related rebound, ERS).^20^ These correlates of movement are typically affected by neurological disease; in people with movement disorders, such as Parkinson’s disease, changes in beta power are related to stage of disease and medication, and a spectral slowing predicts cognitive decline.^21^

The modulation of beta power is not restricted to the motor system, but reflects a broader mechanism for changing states, behaviours and behavioural sets, including behavioural adaptation during motor learning. For example, anticipatory motor preparation to a target stimulus (such as during serial reaction time tasks) is associated with enhanced beta suppression and improved learning.^22–24^ This may reflect motor readiness anticipating the next movement based on prior experience. In people with motor impairments (such as in Parkinson’s disease or after stroke) beta suppression remains limited even after a period of learning, and is related to the reduction in task performance.^23, 25^

Changes in beta power are also associated with inhibitory control.^26^ In people with frontotemporal dementia, clinical manifestations of disinhibition are associated with impaired beta frequency-specific power.^18, 27^ During a response inhibition task (a ‘Go-NoGo’ paradigm), the degree of the beta ERD correlated with carer reports of disinhibition: those with greater disinhibition only successfully prevented a response when there was minimal beta desynchronization, suggesting they maintained a ‘ready state’ to move.^18^ In contrast, in people with amyotrophic lateral sclerosis (ALS), the beta ERD during movement preparation was enhanced, indicating a need for greater desynchronization for effective movement^28^. Notably, beta power underpinning inhibitory control is centred on the right inferior frontal gyrus,^26^ a region that is atrophic in bvFTD and PSP, deficient in neurotransmitter GABA, and linked to inhibitory impairments.^29^ Moreover, this region can respond to pharmacological manipulation: citalopram (a selective serotonin reuptake inhibitor) enhanced the response in the right inferior frontal gyrus in people with bvFTD undertaking a response inhibition task, although there was no concomitant effect on observed behaviour.^27^

We proposed that the impact of FTLD pathologies on beta frequency power is in part related to reduced levels of cortical GABA. GABA directly influences beta power,^30, 31^ and motor responses are dependent on a balance of GABA and Glutamate.^32, 33^ Importantly, people with FTLD disorders who have lower concentrations of GABA are more impulsive.^29^

The vital question is then, whether modifying GABA could restore beta power and consequently improve adaptive behaviour in patients with FTLD-associated disorders. Pharmacological manipulation of GABA levels alters beta power in health and disease. Tiagabine binds selectively and with high affinity to the GABA reuptake transporter GAT1 elevating the level of GABA in the extracellular fluid and synapse.^34^ In health, tiagabine alters beta power widely across the cortex,^35, 36^ and enhances movement related beta ERD/ERS^37^. However, the behavioural consequences of pharmacologically altering beta are unclear; Muthukumaraswamy et al.,^37^ reported no drug induced changes in movement performance in their group of young healthy adults although there were observed changes in beta power. However, restoring GABA in GABA-depleted clinical populations cannot be assumed to be similar to GABA ‘overdose’ in healthy controls.

In this study, we test the relationship between motor performance, motor adaptation, and task-induced beta frequency power. We examined the impact of GABA modulation on performance and beta power using tiagabine 10mg. We included people with behavioural variant frontotemporal dementia (bvFTD) and progressive supranuclear palsy (PSP).^38, 39^ These two conditions exhibit overlapping behavioural symptoms, including cognitive inflexibility, disinhibition, impulsivity and apathy, and impaired social cognition,^39–42^ and both have comparable GABAergic deficits, despite differences in molecular pathology and regional atrophy patterns.^29^

Using a placebo-controlled double-blind randomised cross-over design, we measured movement performance with a novel ‘Controlled Action Response’ (CAR) Task. Magentoencephalography (MEG) measured the changes in spectral power in relation to task performance and to drug. Our predictions were twofold: first we predicted that the bvFTD and PSP participants would show reduced motor adaptation on the task with a concomitant reduction in low frequency power, including loss of beta suppression and rebound. Second, we predicted a differential effect of drug on patients and controls: specifically, that only in the GABA-depleted patient groups would tiagabine restore beta power and improve adaptive movement control, while the control participants were expected to have minimal behavioural changes, despite any observed changes in beta power.

## Materials and methods

### Participants

Twenty-two adults with disorders associated with frontotemporal lobar degeneration were recruited from the specialist frontotemporal dementias clinic at the Cambridge University Hospitals NHS Trust. Eleven had behavioural-variant frontotemporal dementia (bvFTD), and eleven had progressive supranuclear palsy (PSP, Richardson’s syndrome). Diagnoses were made by a consultant neurologist in a multidisciplinary clinic, based on the international consensus clinical diagnostic criteria for probable bvfTD^39, 43^ or probable PSP^38^ including those originally presenting with PSP-F phenotype and progressed to PSP-RS according the MAX-rules criteria for PSP.^44^ People with other PSP phenotypes, other types of dementia, primary progressive aphasias or major psychiatric disorders were not included. A control group of 20 age-matched healthy older adults were recruited from either the volunteer panel of the MRC Cognition and Brain Sciences Unit or the Join Dementia Research register. None had a history of significant neurological or psychiatric illness. Exclusion criteria included current taking of any GABAergic medications, or known adverse reactions to tiagabine or closely related drugs, heart disease or significant cardiac rhythm abnormalities, epilepsy, pregnancy, myasthenia gravis, renal failure. The study was approved by the local Research Ethics Committee and all participants gave written informed consent according to the 1991 Declaration of Helsinki. The Medicines and Healthcare products Regulatory Agency (MHRA) confirmed that the study protocol lies outside the Medicines for Human Use Clinical Trials Regulations 2004 (see also the MHRA clinical trials algorithm).

All participants underwent baseline neuropsychological assessment including the revised Addenbrooke’s cognitive examination (ACE-R)^45^, the Mini-Mental State Examination (MMSE), INECO Frontal Screening Test^46^ the Hayling Sentence Completion Test^47^, frontal assessment battery (FAB)^48^ and the Revised Cambridge Behavioural Inventory (CBI-R) (Wear et al., 2008). Participants with a PSP diagnosis also had a PSP rating scale (PSPRS) examination.^38^ Caregivers completed the revised Cambridge Behavioural Inventory (CBI-R)^49^. One participant with bvFTD and one control could not complete both sessions and were removed from further analyses. Details are summarized in Table 1.

**Table1.** Demographics, cognitive and behaviour test scores. For clinical, cognitive and behavioural tests, best possible scores are shown in parenthesis. Bayesian ANOVAs were used to examine group differences, where there is evidence for a difference between groups (BF > 3) posthoc results are shown for bvFTD vs PSP. Conventional thresholds for Bayes Factors represent evidence in favour of the hypothesis substantial (>3), strong (>10) and very strong (>30). BF < 1 is considered evidence for the Null hypothesis. CBI-R = Revised Cambridge Behavioural Inventory; F = female; M = male; WM = working memory

|  | Controls |  | PSP |  | BvFTD |  | Patients vs Controls | PSP vs bvFTD |
| --- | --- | --- | --- | --- | --- | --- | --- | --- |
|  | Mean / SE |  | Mean / SE |  | Mean / SE |  | BF10 incl | BF10 U |
| Demographics |  |  |  |  |  |  |  |  |
| Group size | 19 |  | 11 |  | 11 |  |  |  |
| Gender | M9 : F10 |  | M7 : F4 |  | M10 : F1 |  |  |  |
| Age | 66.4 | 0.9 | 68.5 | 2.7 | 63.6 | 2.2 | 0.5 |  |
| Education (yrs) | 16.4 | 0.5 | 14.2 | 0.9 | 16.0 | 0.9 | 0.9 |  |
| Handedness | 14.5 | 0.4 | 13.6 | 1.4 | 13.6 | 1.4 | 0.2 |  |
| TGB Blood Values | 141.6 | 9.4 | 145.2 | 22.4 | 130.4 | 15.9 | 0.2 |  |
| Clinical Scales |  |  |  |  |  |  |  |  |
| PSP Rating Scale (80) | ~ | ~ | 35.8 | 3.2 | 10.1 | 1.4 | ~ | 2287 |
| FRS Rating Scale (0/30) | ~ | ~ | 7.7 | 1.76 | 7.1 | 1.1 | ~ | 0.4 |
| Cognition |  |  |  |  |  |  |  |  |
| MMSE (30) | 28.6 | 0.3 | 26.5 | 0.6 | 25.4 | 1.3 | 9.7 | 0.5 |
| ACE-R |  |  |  |  |  |  |  |  |
| Total(100) | 95.4 | 0.9 | 77.9 | 1.7 | 75.3 | 3.6 | 3.4+6 | 0.45 |
| Attention(18) | 17.5 | 0.1 | 16.9 | 0.5 | 16.0 | 0.9 | 0.8 | ~ |
| Memory(26) | 24.1 | 0.7 | 21.6 | 1.2 | 17.5 | 1.6 | 81.3 | 1.5 |
| Verbal Fluency(14) | 13.0 | 0.2 | 4.2 | 0.7 | 5.0 | 0.7 | 1.4+13 | 0.4 |
| Language(26) | 25.4 | 0.2 | 24.1 | 0.5 | 23.2 | 0.9 | 5.5 | 0.5 |
| Visual spatial(16) | 15.4 | 0.9 | 11.1 | 0.9 | 13.5 | 1.0 | 162 | 1.1 |
| INECO |  |  |  |  |  |  |  |  |
| Total (30) | 25.2 | 0.7 | 17.3 | 1.2 | 11.3 | 1.8 | 3.0+7 | 5 |
| WM index (10) | 7.3 | 0.3 | 4.1 | 0.6 | 3.3 | 0.5 | 9.6+4 | 0.5 |
| FABTotal (18) | 17.3 | 0.3 | 11.8 | 0.8 | 11.4 | 1.4 | 1.8 +5 | 0.3 |
| HaylingTest |  |  |  |  |  |  |  |  |
| Scaledscore (23) | 17.9 | 0.3 | 10.0 | 2.0 | 7.8 | 0.9 | 1.5+5 | 0.5 |
| Overallscore (10) | 5.9 | 0.1 | 3.2 | 0.6 | 1.6 | 0.3 | 1.6+7 | 1.7 |
| A + B Converted Error (128) | 4.6 | 1.1 | 17.6 | 4.8 | 39.4 | 5.5 | 5.4+5 | 4.4 |
| **Behaviour** |  |  |  |  |  |  |  |  |
| CBI-R |  |  |  |  |  |  |  |  |
| Total (170) | ~ | ~ | 54.5 | 8.3 | 88.5 | 7.8 | ~ | 6.5 |
| Memory and Orientation (32) | ~ | ~ | 6.9 | 1.3 | 16.1 | 1.6 | ~ | 95.0 |
| Everyday skills (20) | ~ | ~ | 13.3 | 2.1 | 9.6 | 1.5 | ~ | 0.4 |
| Self care (16) | ~ | ~ | 7.4 | 2.0 | 4.0 | 1.4 | ~ | 0.4 |
| Abnormal behaviour (14) | ~ | ~ | 3.7 | 0.7 | 13.0 | 2.0 | ~ | 94.2 |
| Mood (16) | ~ | ~ | 1.8 | 0.5 | 5.5 | 1.2 | ~ | 6.2 |
| Beliefs (12) | ~ | ~ | 0.2 | 0.1 | 0.8 | 0.4 | ~ | 0.9 |
| Eating habits (16) | ~ | ~ | 4.5 | 1.5 | 10.2 | 1.6 | ~ | 3.3 |
| Sleep (8) | ~ | ~ | 3.5 | 0.8 | 3.5 | 0.7 | ~ | 0.3 |
| Stereotypic and motor (16) | ~ | ~ | 3.2 | 1.4 | 11.5 | 1.5 | ~ | 40.2 |
| Motivation (20) | ~ | ~ | 10.2 | 2.0 | 14.1 | 1.7 | ~ | 0.8 |

### Experimental design

The study was registered with ISRCTN registry (ISRCTN10616794). All participants were entered into a randomized double-blind placebo-controlled crossover design. Two sessions were conducted approximately two weeks apart, in which participants were given a single dose of either 10 mg oral tablet of tiagabine or matched placebo. The tiagabine or placebo were dispensed prior to the study into bottles labelled with participant and visit number by the hospital pharmacist. The randomisation order was permuted in sequential blocks of six participants, and known only by the dispensing pharmacist. The tablets were administered by the Cambridge Clinical Research Centre nurses each visit. The study researchers involved in the analysis were not involved with administering medication to the patient, and were not present in the room when the tablets were taken. Venus blood samples were taken approximately 105 minutes after drug administration, immediately before the MEG recording, close in time to the estimated time of peak plasma levels and CNS penetration^50^. Mean plasma levels were measured by a specific validated high performance chromatography. A comparison across controls, PSP and bvFTD groups showed evidence of equivalence for the level of tiagabine in participant serum (Bayesian Analysis of Variance, BF10=0.2, in favour of no group difference). Power analyses for frequentist tests showed a power >0.8 to detect a medium effect size in a repeated measures ANOVA, for a group by drug interaction (effect size F = 0.25, 3×2 factor, N=42; G*Power software version 3.1.9.2; Heinrich-Heine-Universität Düsseldorf, Germany).

### Task paradigm

We used a novel computerised visuomotor task to measure the ability of participants to control an on-screen cursor, known as the CAR task (controlled action response). The cursor was in the shape of a car (visual angle = 1.2°), presented 5.5° to the right of centre on the screen and a target box (visual angle = 1.6°) presented 5.5° to the left. Participants used a MEG compatible joystick and were asked to ‘move the car cursor into the target parking space as quickly as possible’. The joystick acted as a throttle to accelerate or decelerate the speed of the car. To promote an adaptive behavioural response, the ‘gear’ of the car (i.e. the joystick gain) was randomly changed between short blocks of trials which determined the output speed of the car. Four gear types were used in which the cars maximum speed was either: 0.58, 0.33, 0.21, 0.15 pixels per ms. Trials with the same ‘gear’ were repeated in succession for short blocks of 4 to 8 trials, before changing to a different ‘gear’ selected at random. Trial order was permuted such that all participants received the same trials, but in different orders. There were 384 trials in total, 96 of each gear speed. For each trial, the car appeared on the screen in the same start position, cueing the beginning of the trial. The car was always moved from right to left on a fixed horizontal plane, due to the greater ease of an inward wrist flexion. When the car was stopped inside the target box positive feedback (a green tick) was provided, whereas negative feedback (a red cross) was provided if the car was stopped outside the box, or if the car was not stopped within 2000ms from onset time. All feedback was presented centrally on the screen for 200ms. The interstimulus interval was randomly varied between 600-1000ms. Trials in which the car was not moved and trials in which the car was accelerated to the edge of the screen were not included in the analyses, as in these trials no control of the car was evident. The mean number of trials included in the behavioural analyses for the control group was 384 (SD = 0) and for the patient group 357 (SD = 66). Three of those in the patient group (one with bvFTD, and two with PSP) stopped the task before the last block, but had sufficient trials to include in the analyses (mean = 143, SD = 29).

### Behavioural Analyses

Behavioural analyses examined the following performance features: accuracy (position of the car at the end of the trial), time of movement onset, the total duration of movement (from time of first movement to stop), and ‘mean-error’ calculated as the mean position of the car sampled every 60ms throughout the 2000ms epoch. The mean-error provides a score that reflects both accuracy and movement duration. Mean differences of these behavioural measures were examined with repeated measures analysis of variance, supported with evidence from Bayesian analyses of variance, conducted in JASP (Version 0.10.1, JASP Team, 2020; jasp-stats.org). The ANOVA reports group differences (Controls, vs bvFTD and PSP), drug effects (tiagabine vs placebo) and the interaction between group and drug. For the Bayesian analyses thresholds for interpretation are Bayes factors >3, >10, >30, and >100 representing weak, moderate, strong and very strong evidence, respectively.^51^

To examine behavioural adaptation or learning over trials, we used a linear mixed model to estimate the change in mean-error across trials, using the *fitlme* function in Matlab R2018a. The individual mean-errors from each participant and each session were concatenated and outliers of more than three median absolute deviations were removed, the data were then z-transformed. The assumptions of a normal distribution were checked by means of Q–Q plots and Shapiro-Wilk and Kolmogorov-Smirnov tests. The model included the fixed factors: trial index (1 to 384 as a continuous variable), short-block trial index (1 to 4-8 as a continuous variable), drug session (placebo/tiagabine as a categorical variable) and group (controls/PSP/bvFTD as a categorical variable). Random effects included the ‘gear’ (1-4) and participant number. An ANOVA using the Satterthwaite correction estimated effects of trial index, which would indicate whether there was an improvement in performance as the trials progressed, and differences and interactions between group and drug.

For each individual participant a slope of their estimated mean-error across trials was calculated using a 1^st^ order polynomial fit, which represented a ‘learning slope’. To examine the relationship between cognitive ability and task behaviour in the patients, mean-errors and the individual learning slopes were correlated with cognitive tests of frontal function (INECO and Hayling tests) and general cognitive ability (ACE-R).

### Structural MRI

A T_1_-weighted structural image (magnetization prepared 2 rapid acquisition gradient echoes, MP2RAGE) was obtained from each subject at 7 T on a Siemens TERRA system (Siemens Healthineers, Erlangen, Germany) with 32 channel headcoil. Acquisition parameters were: 0.75 mm isotropic voxels, TE = 1.99 ms, TR = 4300 ms, inversion times = 840 ms/2370 ms). Two patients were unsuitable for 7 T and underwent 3 T scanning on a Siemens PRIMSA system (Siemens PRIMSA MPRAGE; 1.1 mm isotropic voxels TE = 2.9 ms, TR = 2000 ms). Two controls declined MRI. The individual structural scan was used to co-register the MEG data to enable subject-specific modelling of the lead field for the beamformer analyses. For the participants in whom the MRI was unavailable the default template was used instead.

### MEG Data

#### Preprocessing

MEG data were acquired at the MRC Cognition and Brain Sciences Unit, using a 306- channel Vectorview MEG system (Elekta Neuromag), which contains two orthogonal planar gradiometers and one magnetometer at each of 102 positions. Data were recorded continuously at 1 kHz in a magnetically-shielded room. Five head position indicator (HPI) coils were used to monitor head position. Vertical and horizontal electrooculograms were recorded using paired EOG electrodes. The 3D locations of the HPI coils, over 100 ‘head points’ across the scalp, and three anatomical fiducials (the nasion and left and right pre-auricular points), were recorded using a 3D digitizer (Fastrak Polhemus Inc.).

The raw MEG data were initially preprocessed using MaxFilter software (version 2.2, Elekta-Neuromag). Bad channels were detected by MaxFilter’s ’autobad’ option (and defined as bad if bad in more than 5% of recording) and reconstructed by MaxFilter. The raw continuous data were cleaned using the spatiotemporal extension of the signal separation algorithm (tSSS).^52^ The origin of the SSS expansion was determined by fitting a sphere to all digitized head points. The data were corrected for head motion at least every 1s. Data sets were realigned into a default head space using the Maxfilter ‘trans’ function. Further preprocessing and data analysis used MATLAB (The MathWorks, Natick, MA) and SPM12 (SPM, Wellcome Trust Centre for Neuroimaging, London).

Data were downsampled to 250 Hz and high pass filtered at 0.1 Hz. Independent component analyses were used to automatically detect and remove artefacts related to eye movements and blinks, performed in EEGlab (Swartz Center for Computational Neuroscience, University of California San Diego). Epochs of 2500 ms were extracted (−500 ms to 2000 ms) time-locked to the onset of the car cue. Epochs containing artefacts were rejected if the amplitudes exceeded thresholds of 2500 fT for magnetometers and 900 fT for gradiometers. One control and one patient with PSP were removed from further MEG analyses due to excessive noise in the MEG data.

After artefact rejection, and exclusion of trials for behaviour as described above, the mean number of trials included in the MEG analysis for the control group was 365.9 (SE = 5.5) and for the patient group 295.2 (SE = 21.0). The preprocessed data were then entered into the time frequency analysis and source space analysis described below.

#### Time-frequency in sensor space

Time-frequency power spectra were computed for frequency bands between 6–44 Hz across each whole epoch using Morlet wavelets with a factor of 7, baseline corrected using a log ratio of power and scaled to dB. Time-frequency images for each separate trial were created and smoothed with an 8mm kernel. Statistical analyses were performed on these images in a 2-step process. First, the trial time-frequency images were entered into participant specific ANOVAs, together with mean corrected covariates of trial behaviour, including: the start of the response in ms, the duration of the response in ms, the mean accuracy of the response, the trial index of the short-blocks (1:8), and the trial index across the whole task (1:384). Second, to examine differences and interactions between the drug conditions and groups, the Beta images for each covariate from the first step, were entered into separate 3×2 random effects ANOVAs. The statistical maps were thresholded with a cluster-based family-wise error (FWE) correction *P* < 0.05 (after *P* < 0.001 voxel-wise height threshold).

#### Source reconstruction

Source reconstruction was performed in SPM12. The preprocessed MEG data were coregistered to each participant’s individual anatomical T1-weighted MRI image using the digitised fiducial and head points. The forward model (lead field) was estimated from a single shell cortical mesh. Inverse source reconstruction was computed using the SPM12 Linearly Constrained Minimum Variance (LCMV) beamformer^53^, for the beta (12-30 Hz) band. Model fit was good: R^2^ model fit in patients: *M =* 91.49; *SD =* 6.56; R2 model fit in controls: *M =* 92.49; *SD* = 4.62.

Images from the LCMV were computed for each subject, and included the mean of all trials across three time windows of interest, with a width of 400 ms, centred on 750 ms, 1250 ms and 1750 ms after stimulus onset to encompass the desynchronisation and rebound, and also baseline images for each trial type (−500 to -100 ms).

Beamformer source images for all participants were entered into 3×2×2 random effects ANOVAs, including participant group (Control, PSP, bvFTD) , drug session (tiagabine or placebo), and source image (baseline and task window). Three ANOVAs were conducted for each of the time windows of interest. For each ANOVA, two sets of contrasts were estimated: First, trial-modulated beta power examined the magnitude of change across the window of interest from baseline (task window - baseline) in order to examine the extent of ERD/ERS. Second, task-related beta power examined the absolute mean difference in beta power without a baseline correction in order to examine group and drug differences within the context of the whole task. Statistical estimations tested differences between the three participant groups and interactions with drug condition. The statistical maps were thresholded with a cluster-based family-wise error (FWE) correction *P* < 0.05 (after *P* < 0.001 voxel-wise height threshold).

## Results

### Behaviour

The mean responses for the behavioural measures of accuracy, mean-error, and movement times are presented in Figure 1 A-D. The repeated measures ANOVA for accuracy confirmed a significant effect of group (F_(1,37)_=3.6, *P<*0.05; BF = 1.6); drug (F_(1,37)_=11.44, *P<*0.05; BF_10_ = 25160) and a group x drug interaction (F_(2,37)_=5.3, *P<*0.05; BF_10_ = 1.447e ^+6^), indicating that the patients tended to undershoot (i.e. the car was stopped before the target box), and this was increased in the drug condition. Post-hoc tests showed this difference was greatest between controls and PSP participants (t = 2.6, *P<*0.05; BF_10_ = 2.850e +21). There were no differences in the mean-error measures. For the movement timings, there was a significant difference between groups in movement duration (F_(2,37)_=5.9, *P<*0.05; BF = 8.6). The post hoc comparisons showed the PSP group to have longer response durations compared to the other two groups (post hoc comparisons: Controls vs PSP, *t*=3.23, *P<*0.05, BF_10_ =1.1^+71^ ; bvFTD vs PSP, *t*=2.8, *P<*0.05, BF_10_ =2.3^+51^). There was no evidence of drug or group effects on any other measures. Across the patient groups, there was a significant correlation of mean-error with ACE-R total (Pearson’s R = -0.43, *P<* 0.05) (Figure 1I), indicating that those with better general cognition had lower Mean-error, although there was no significant relationship with the INECO (Pearson’s R = -0.29, ns, Figure 1J).

**Figure 1.**
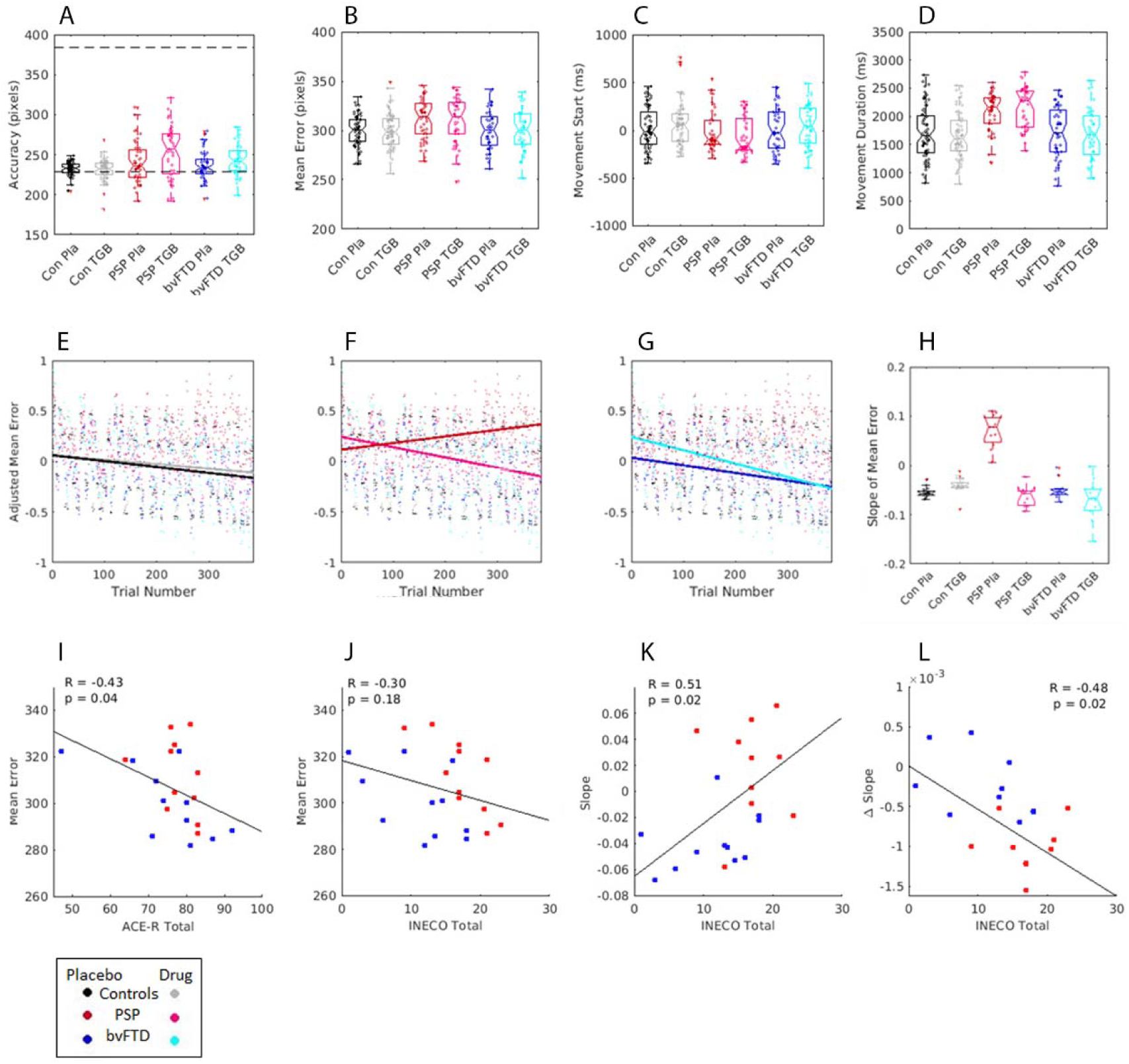
Behavioural task results. Plots A-D show bar plots of the individual’s mean scores for accuracy, mean-error, movement start time and movement duration time. In plot A, the dotted line represents the cue start point and target end point. Plots E-G display the adjusted Z-score of mean error over trials, estimated from the linear mixed effects regression, a positive slope shows an increase in mean-error over trials, where as a negative slope shows a decrease in mean-error over trials. H is a bar plot of the slope of learning for each group, estimated from the polynomial coefficients of slope for mean-error over trials. Plots I and J are scatter plots of the relationship between mean-error on placebo and cognitive scores for ACE-R and INECO. Plot K shows the relationship between the INECO cognitive test and the slope of error over trials on placebo, L plots the change in slope when on tigabine. For all plots, PSP is in red, bvFTD is in blue and controls are in black.

We measured the participants’ performance over the course of the session, as they learned to adapt to the changing gain of the joystick (the changing “gear”). A mixed effects linear regression estimated the learning effects as the slope of the change in mean-error over trials (Figure 1E-G, Mean ‘Learning slope’ for each group is presented in Figure 1H). The model fit was good, R^2^ adjusted = 0.4. ANOVA revealed a significant main effect of trial index, with a decrease in mean-error progressing over the trials (*F*_1,22238_= 20, *P* <0.01), but no significant main effect of short-block trial index, drug session or group. There were significant interactions between trial index and group (*F*_1,22244_= 6.6, *P* <0.05), and a significant interaction of trial index, drug and group (*F*_1,22239_= 6.7, *P* <0.001).

Tiagabine improved motor learning in the patient groups. Specifically, post hoc tests of the fixed effects coefficients for these interactions revealed the PSP group have impaired motor adaptation compared to controls (F_(1,21717)_=12.3, *P<*0.05) and the bvFTD group (F_(1,21717)_=6.5, *P<*0.05), while there was no significant difference between the bvFTD group and controls (F_(1,21717)_=0.5, ns ). To test for the effect of drug on learning, in patients versus controls, a post-hoc test of the three-way interaction (trial x drug x group) revealed that tiagabine improved the mean-error over trials for the PSP participants (F_(1,21717)_=8.8, *P<*0.05) with marginal improvements in the bvFTD group (F_(1,21717)_=3.4, p=0.06); and no significant difference between the bvFTD and PSP groups (F_(1,21717)_=1.0, ns);

Tiagabine benefited behavioural adaptation in those who had relatively preserved cognition. Specifically, among patients there was a significant relationship between the learning slope in the placebo condition and the individuals’ INECO (Pearson’s R = 0.51, *P<* 0.05, Figure 1K), but not with their ACE-R (Pearson’s R = 0.17, *P<* 0.05); and a significant relationship between the change in learning slope when on tiagabine (learning coefficients of drug condition – placebo condition) and INECO score (Pearson’s R=-0.48, *P<*0.05, Figure 1L). These correlations indicate that those who have lower INECO scores show a greater change in behaviour over trials, however it is those patients with relatively higher INECO scores who benefit more from tiagabine, and show greater improvements over trials when on drug. There was no significant relationship between learning with general cognition, as measured by ACE-R total score (Pearson’s R=-0.17, ns).

### Time-Frequency in sensor space

The mean time-frequency spectra for all trials is depicted in Figure 2. The results confirm the predicted event related desynchronization (ERD), which is prolonged throughout the movement followed by a rebound late in the epoch, reflecting the movement onset and offset. The ANOVA comparing the three groups (Controls, PSP, bvFTD, Figure 2G) revealed significant loss of Beta power during the ERD window (peak at 22-24Hz, 964-1208 ms) and reduced alpha/beta power at trial onset in the patient groups, particularly evident in the PSP group (peak 12Hz 208ms, Figure 2H). There were no significant global differences on tiagabine, nor an interaction between group and drug.

**Figure 2.**
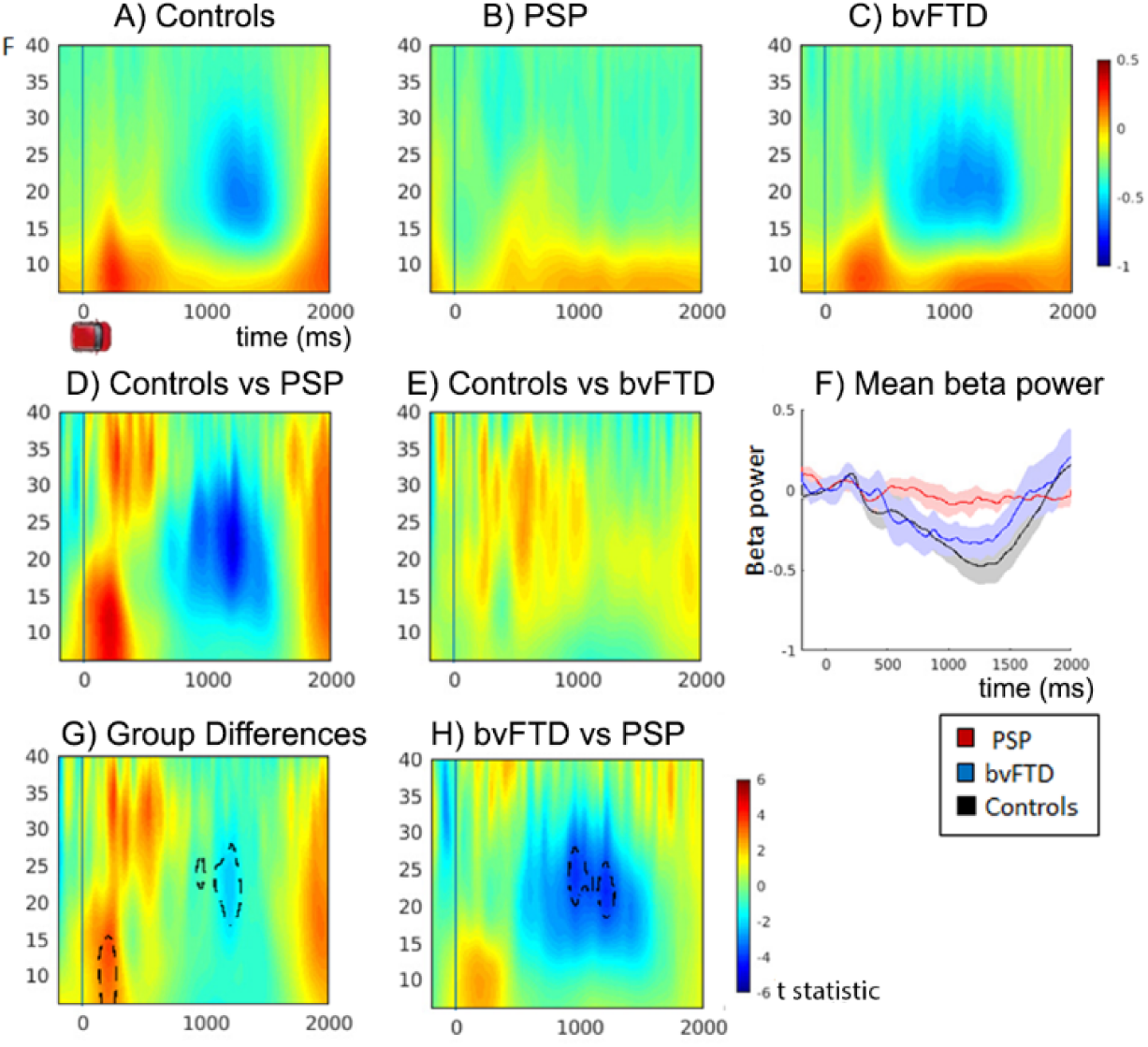
Time Frequency plots showing changes in frequency power on placebo. Plots A-C show mean time-frequency power over all trials for Controls, PSP and bvFTD groups, increased power is displayed in red and decreased power in blue. Epochs are time locked to the onset of the car cue, time point 0. D & E) Difference plot of controls vs PSP and controls vs bvFTD, plots are the *t* statistic of the difference. F) Line plot of mean beta power for each group across time, with CI. G) Power differences between all groups, and H) power differences between bvFTD vs PSP. Plots D,E,G and H plot the *t* statistic of the differences. Significant clusters from the F test of group differences are traced with black dotted lines (p < 0.05 fwe cluster wise correction after p<0.001 voxel-wise threshold).

To investigate the relationship between spectral power and task behaviour, and to examine between group and drug effects, two-factor ANOVAs (group x drug) including behavioural covariates were conducted. The behavioural covariates included: trial progression, mean-error, movement start time and duration. There were no significant effects of the short in-block trials, in behaviour or in the MEG, and these are not further reported.

The mean change in frequency power over trials is shown in Figure 3 (for separate group means on drug and placebo, see supplementary information, Figure S1). With trial progression (Figure 3a) spectral power was enhanced: revealing an increase in beta power after trial onset, in line with the joystick being held in a steady position (peak cluster at 20 Hz, 496 ms), followed by an enhanced ERD as the car cursor was positioned into the target box (peak cluster at 16 Hz, 936 ms).

**Figure 3.**
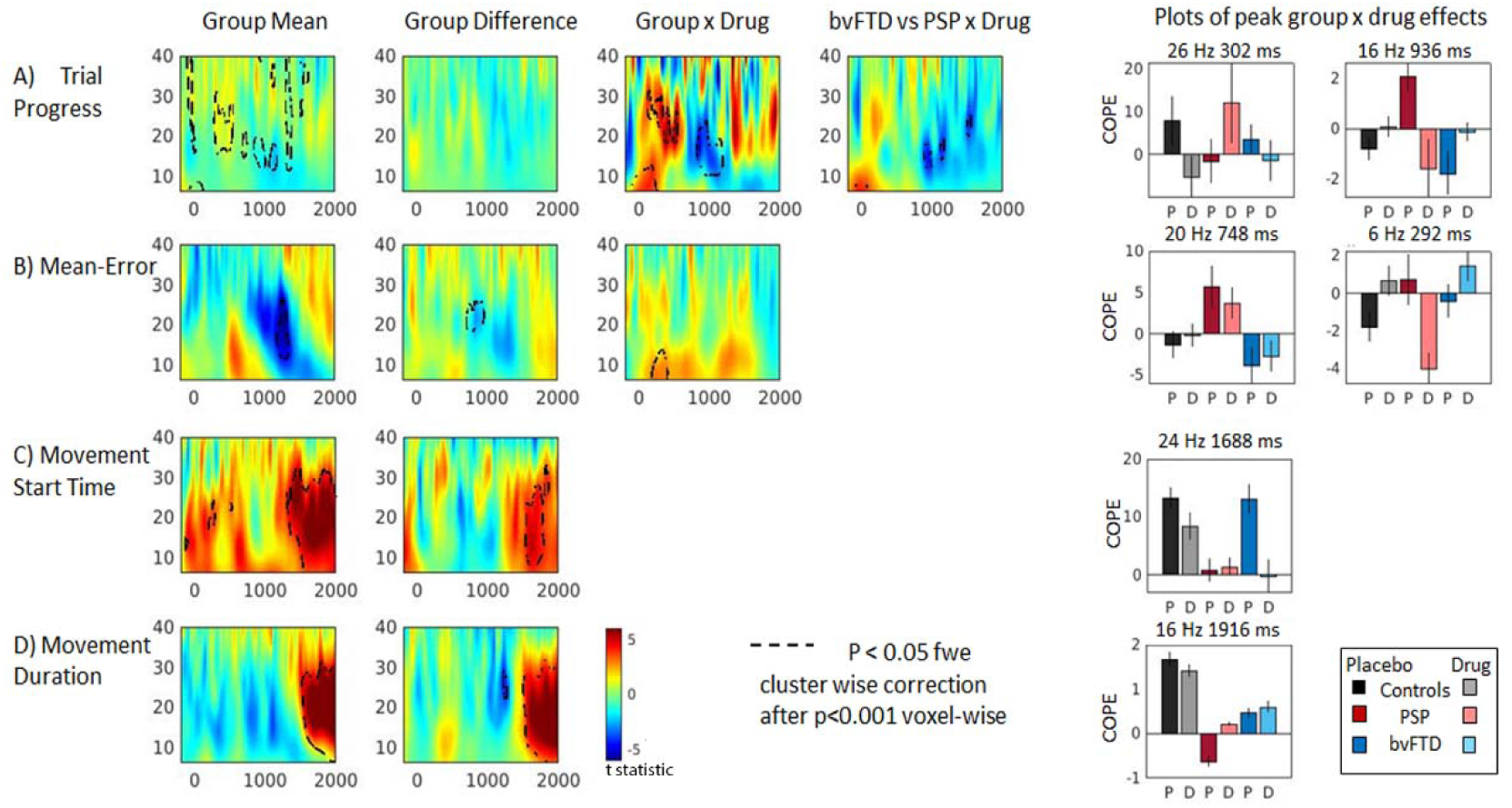
Time Frequency plots showing the relationship between changes in frequency power and task or behavioural covariates. Plots are the t statistic of group means or differences. Black dotted lines highlight significant clusters (p<0.05, fwe cluster wise correction after p<0.001 voxel-wise threshold). Group means show mean across all groups; Group difference shows impaired frequency power in one or more patient groups compared to controls; Group x Drug and bvFTD vs PSP x drug, shows where drug modulates frequency power in one group compared to another. A) Changes in power with trial progression. Red shows increases in power as trials progress, and blue shows decreases. B) Mean–error: power decreases (blue) and increases (red) associated with lower mean-error (better performance). C & D: beta power increases (in red) indicate the ERS which is greater with an earlier movement start time, and shorter movement durations. E) Bar plots show direction and magnitude of peaks within significant clusters of the tests of interaction. Placebo (P) and Drug (D). Error bars are standard error.

In the two patient groups, the relationship between trial progression and beta power differed from the controls in a significant interaction with tiagabine. Two main clusters in the beta band (Figure 3A Group x Drug peaks 26 Hz 302 ms, 16 Hz 936 ms) and an early theta/alpha cluster (Figure 3A Group x Drug peak 6 Hz, 8 ms) were differentially affected by drug. In PSP on placebo, beta power increased in these clusters with trial progression, leading to a reduced ERD in later trials, but on tiagabine, similar to the controls, there was greater desynchronisation with trial progression during the ERD window. In bvFTD, on placebo there was enhanced desynchronisation with trial progression, but on tiagabine beta power was elevated, minimising the ERD (Figure 3A bvFTD x PSP peak 16 Hz 936 ms)

Lower mean-error was associated with a greater ERD after 1000 ms (Figure 3B). There was a significant difference between the three groups (Figure 3B Group difference peak 20 Hz 748 ms), with PSP patients showing elevated beta power with lower mean-error in this cluster, compared to the control and bvFTD groups in whom an enhanced ERD was associated with lower error. The interaction with tiagabine was in the early theta/alpha window (Figure 3B Group x Drug peak 6 Hz 292 ms), which was elevated by the drug in controls and bvFTD, but reduced in PSP.

Earlier movement start times and shorter movement durations were associated with a significantly enhanced beta rebound after 1500 ms, (Figure 3 C & D, movement start peak at 20 Hz 1668 ms; movement duration peak at 18 Hz 1924 ms). The beta rebound was diminished in the patient groups, with peak differences after 1500 ms (movement start peak at 24 Hz 1688 ms; movement duration peak at 16 Hz 1916 ms).

### Source reconstruction of disease and drug effects

The source reconstruction aimed to identify regions in which beta power was being significantly modulated within trial, and within the context of the whole task. Within trial beta power was examined by measuring the magnitude of change between trial baseline (-500 to -100 ms) and three 400 ms windows centred on 750, 1250 and 1750 ms. Task related beta power examined the absolute mean difference in beta power between the groups, using the same three windows centred on 750, 1250 and 1750 ms .

The trial-related beta desynchronization was localised to bilateral pre and post central gyrus and right superior parietal regions in all groups (Figure S2 A-B), and increases in beta power were localised to anterior frontal and temporal regions. During the rebound window (Figure S2C) increases in beta power were also localised to left motor cortex.

There were no significant differences between groups in the localisation of the desynchronization within motor cortex.

Both groups of patients, compared to controls, had reduced beta power in occipital and temporal regions (1250ms window, Figure S2 G), and a loss of rebound localised to left precentral gyrus (1750 ms window, Figure S2H, cluster peak -16 0 44). There was a small drug by group interaction in the left insula (Figure S2G in violet, peak of cluster - 28 -18 22), with tiagabine enhancing beta power in the PSP group, and reducing beta power in the bvFTD group.

The critical comparison is of task related beta power differences between the groups as shown in Figure 4, (for the same three time windows as the above analysis). Relative to the placebo session, the beta power in right inferior frontal cortex was reduced (i.e. improved) by tiagabine in the patient groups, but increased (abnormally reduced ERD) in the control group. This confirms that the beta ERD in the right IFG was enhanced in patients by the drug (Figure 4 A-C, bar plot shows peak at 32 28 14). Differences between the two patient groups showed that bvFTD patients had greater beta power in right superior parietal cortex compared to the PSP group, but there was no effect of drug (Figure 4 D-F).

**Figure 4.**
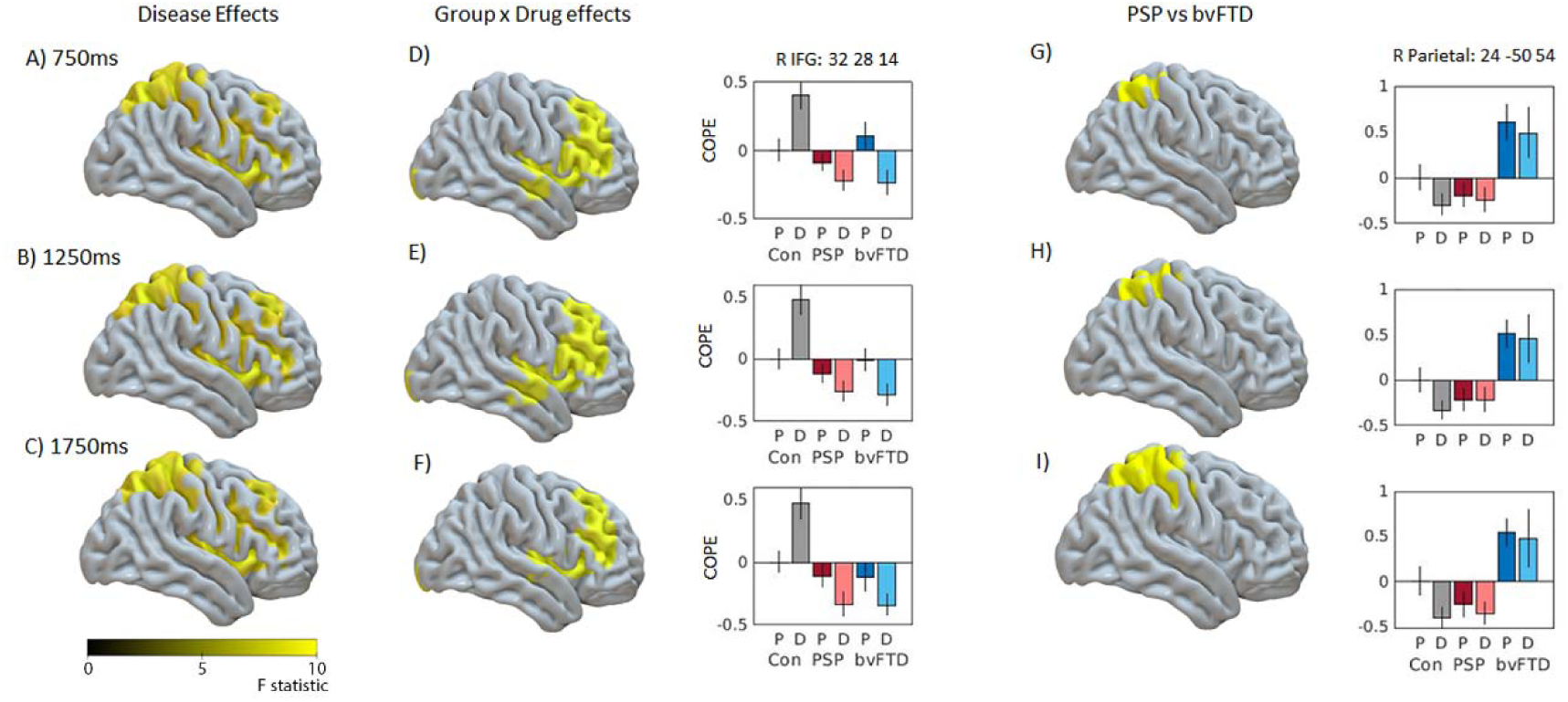
LCMV beamformer source reconstruction of task related beta power. Figures A-C show mean difference between the three groups in right prefrontal and right parietal cortex, and D-F show group by drug interactions in right prefrontal cortex. Images show mean source power of three time windows, centred on 750, 1250, 1750 ms. Clusters shown are familywise error corrected (*P* < 0.05 after *P* < 0.001 voxelwise uncorrected threshold). Bar plots show peaks of significant clusters of drug interaction within right inferior frontal gyrus (peak xyz: 32 28 14). Figures G-I show differences between PSP and bvFTD groups in right parietal cortex. Bar plots show peaks of significant clusters in right posterior parietal cortex (peak xyz = 24 -50 54). Data in bar plots are plotted relative to controls placebo session, to display the direction of beta change. X axis is for each group on placebo (P) and Drug (D), Y axis is contrast of parameter estimate, COPE. Error bars are standard error.

## Discussion

The main outcomes of this study are that (i) one can pharmacologically restore spectral power in people with bvFTD and PSP by treating the GABAergic deficit, and (ii) the drug-induced changes in beta power are accompanied by improved adaptive control of behaviour. Tiagabine improved the behavioural performance of people with bvFTD and PSP over trials, and this improvement was concomitant with changes in beta power, localised to prefrontal cortex. We interpret the response to drug as an improvement in motor adaptation driven by the prefrontal motor-control network. Importantly, the drug had differential neurophysiological effects on the two patient groups: in those with PSP, tiagabine gradually enhanced the beta event related desynchronization (ERD) over trials, while in those with bvFTD tiagabine reduced the ERD. This suggests baseline-dependency by diagnosis in neurophysiological responses to the pharmacological challenge. There was also baseline dependency by cognition, in the effect of tiagabine on task performance. Consistent with previous studies of tiagabine,^37^ there were no observable effects of tiagabine on the behavioural measures of the healthy control group, who have age-related normal baseline GABA.

The close relationship between movement and beta power suppression/rebound is well described.^20^ However, the modulation of beta power is more than a simple neurophysiological index of movement. It has been linked with cognitive and behavioural responses, including response inhibition, and motor learning and adaptation.^54^ An enhancement of beta suppression occurs when the sequence of temporal and spatial cues for a motor response are predictable, facilitating an improvement in performance.^22^ Accordingly, in our control group better performance and trial progression were also associated with enhanced beta suppression, with the opposite observed in the patient groups. This deficit was clearly evident in those with PSP on placebo, in whom performance became worse and beta suppression diminished with trial progression.

Beta desynchronisation and rebound during each epoched trial is typically interpreted as a temporally sustained oscillatory power modulation. However variations in sensorimotor beta power also reflect transient (non-oscillatory) bursting activity.^55, 56^ Mechanistic models of beta power generation reveal that beta-range bursting activity arises within a cortical microcircuit of pyramidal neurons and inhibitory interneurons,^57, 58^ that can generate beta-bursts via a confluence of deep and superficial synaptic drives with laminar specific inhibition.^59, 60^ The effect of bvFTD and PSP on beta power may therefore reflect the specific laminar and inhibitory-synaptic consequences of disease.

In health, the connectivity strength of recurrent inhibitory and excitatory connections are driven by the individual participants’ GABA and Glutamate, as measured by MR spectroscopy.^61^ In patients, attenuated beta bursts may arise from impaired neurotransmission that interrupts the laminar dynamics of cortical microcircuits. Adams et al., have shown that in both PSP and bvFTD the deep inhibitory intrinsic connections are impaired compared to controls, and can be restored by tiagabine.^62^ Notably, Adams et al also observed a differential effect of tiagabine between the PSP and bvFTD groups at the phasic inhibition of the stellate cells – the PSP group had higher inhibition reduced by tiagabine, and the opposite pattern occurred for those with bvFTD. Adams et al., interpret this as a function of cellular loss, with bvFTD causing greater cell loss and atrophy, and consequently tiagabine has a differential effect. Here, we extend that interpretation and suggest that the response to drug may be dynamic, at least in the context of a motor learning paradigm; and that baseline state for movement, in terms of relative beta power changes needed for movement (i.e. ERD), determines the response to drug.

Previously we have shown that participants with bvFTD have a reduced beta ERD even for successful motor responses during a Go-NoGo task.^18^ Critically the NoGo trials, which required the prepotent ‘Go’ response to be withheld, were more successful only when the ERD was minimal, and this was more evident in those with higher scores of clinical disinhibition.^18^ We interpreted this as the participants being in a ‘ready state’ to move, and any ERD prevented a change in neural state to one of motor inhibition. A reduced modulation of beta power is also observed in participants with movement disorders such as Parkinson’s disease or ALS, but in contrast this has been related to akinesia and movement impediment because a high baseline beta power is maintained.^21, 63^ Pharmacological modulation of the GABA system can enhance movement in those with movement disorders, including PSP and PD.^9, 64^ Thus, enhancing GABA could facilitate movement by a dynamic modulation of beta power which depends on the starting baseline of the sensorimotor network.

The fuction of beta power modulation in the context of motor learning, and the critical role of inhibitory interneurons, is explicable within the Active Inference framework for movement preparation and execution.^65–67^ This approach considers movement to be generated via predictions about sensory input, rather than by direct ‘forward’ commands. These predictions arise from generative models which represent the predicted sensory consequences of movement. With effective motor learning or adaptation, the difference between the predicted and perceived sensory input (i.e. the prediction error) is reduced: achieved by updating the predictions or by actively changing the sensory input. The extent of the adjustments are dependent on the uncertainty, or precision, of the sensorimotor predictions and the sensory input. Several studies indicate that the changes in sensorimotor beta power inversely index the precision of the sensorimotor system. ^68–71^ In accordance with this theory, an increase in the ERD/ ERS would index successful learning, and a failure to learn and adapt behaviour would result in minimal changes to beta power. Indeed, when beta suppression is impaired in movement disorders like Parkinson’s disease, sequence acquisition is concurrently impaired^23^. Comparably, this is the pattern we observe in those with PSP, in whom movement and motor learning is impaired and beta power changes are minimal, suggesting impaired sensorimotor precision leading to reduced updating of the forward model to improve performance over trials. The source localisation revealed a prefrontal sensitivity to the effects of tiagabine in patients, suggestive of a more cognitive frontal process involved in motor learning for this task. We speculate that the changes in beta power in prefrontal regions associated with motor learning represents the impairment in sensorimotor precision as a result of FTLD, subject to GABAergic modulation.

There are several limitations to this study. The use of single dose tiagabine in this study is purely for research purposes on the impact to spectral power, with the aim to improve our understanding of the mechanisms of disease. It was not assessed as a clinical treatment or tested against clinical outcome measures, although the current results would support the case for such trials in the future. We included people with PSP and bvFTD, based on clinical diagnostic criteria. Clinico-pathological correlations for PSP-Richardson’s syndrome are very high, to a 4R-tauopathy in ∼95% of cases in our brain bank series. For bvFTD, an FTLD pathology is also identified in ∼95% of cases, albeit with either Tau or TDP43-pathologies. We study these disorders jointly, despite differences in molecular pathology, because of the well-established similarities in cortical physiology, neurotransmitter deficits, and cognitive/behavioural profiles.^17, 38, 42^ The group sizes in this study are relatively small, which raises the question of power. However, we predicted medium to large effect sizes for which our groups size was adequate to achieve >80% power (frequentist tests) and note that the Bayesian analyses had sufficient precision in the data to support inferences. In terms of the pharmacology, tiagabine is not selective by brain region, and it cannot selectively target the most GABA deficient regions in the patients. We did not quantify baseline levels of GABA (by spectroscopy), or the density and distribution of GABA_A_ receptors (by ^11^C-flumazenil PET), which may influence an individual’s response to drug.^29, 36, 72^ While we cannot show the GABA deficit of the specific participants in our study, others have shown a reduction in frontal cortical GABA in people with PSP and bvFTD, ^4, 29^ in proportion to cognition (Perry et al., 2022, SI Fig. 3^12^). The pharmacological effects on beta oscillations may be drug-specific, and we note for example that propofol, a GABAa receptor modulator and agonist, does not have comparable effects on movement related beta oscillations.^73^

## Conclusion

Restoring selective deficits in neurotransmission is a potential means to improve behavioural symptoms in those with bvFTD and PSP, analogous in rational to dopaminergic therapy in Parkinson’s disease. The concomitant neurophysiological and behavioural changes in response to tiagabine is important for drug development for symptomatic treatment in three ways. First, beneficial effects of drug were evident regardless of degree of atrophy in task-related brain regions. Second, the drug response in patients differed from the response in controls, indicative of a baseline dependency. Third, the drug’s effects were not observed within the global mean behaviour, but in the change in behaviour and neurophysiology with learning. This is suggestive of a GABA-dependent beta related mechanism which allowed participants to appropriately adapt and modify behaviour accordingly with experience. We hope this experimental medicines study will inform the design and execution of much needed symptomatic clinical trials to help people affected by disorders associated with frontotemporal lobar degeneration.

## Data Availability

The data that support the results of this study will be available from the corresponding author, upon reasonable request for academic (non-commercial) purposes, subject to restrictions required to preserve participant confidentiality. A data transfer agreement may be required.

## Data availability

The MEG data preprocessing pipeline is available at https://github.com/LauraHughes2024/MEG2024. The data that support the results of this study will be available from the corresponding author, upon reasonable request for academic (non-commercial) purposes, subject to restrictions required to preserve participant confidentiality. A data transfer agreement may be required.

## Acknowledgements and Funding

This work was primarily funded by the Wellcome Trust (220258), with additional support from the Medical Research Council (MC_UU_00030/14; MR/T033371/1) and the NIHR Cambridge Biomedical Research Centre (NIHR203312), and carried out at/ the NIHR Cambridge Clinical Research Facility. This work was co-funded by the Holt Fellowship, Association of British Neurologists and Patrick Berthoud Trust. We thank the PSP Association & FTD Support Group for raising awareness of the study. The views expressed are those of the authors and not necessarily those of the NIHR or the Department of Health and Social Care. For the purpose of open access, the authors have applied a CC BY public copyright licence to any Author Accepted Manuscript version arising from this submission.

## Supplementary material

Supplementary material is available online

